# Clinical care site data integration reveals heterogeneity in EHR phenotyping and healthcare utilization patterns

**DOI:** 10.64898/2026.06.21.26356156

**Authors:** John P. Shelley, Allison M. Lake, Julia M. Sealock, Thomas E. Ueland, Josh F. Peterson, Lea K. Davis, Jonathan D. Mosley

**Affiliations:** Department of Biomedical Informatics, Vanderbilt University Medical Center, Nashville, TN, USA; Vanderbilt University School of Medicine, Nashville, TN, USA; Analytic and Translational Genetics Unit, Massachusetts General Hospital, Boston, MA, USA; Stanley Center for Psychiatric Research, Broad Institute of MIT and Harvard Cambridge, MA, USA; Division of Data-Driven and Digital Medicine, Department of Medicine, Icahn School of Medicine at Mount Sinai, New York, NY, USA; Department of Medicine, University of Texas Southwestern Medical Center, Dallas, TX, USA

**Keywords:** electronic health records, phenotyping, polygenic risk score, healthcare utilization

## Abstract

**Objective:** Genomic research using electronic health record (EHR)-linked biobanks is influenced by heterogeneity in the clinical settings (care sites) where encounters occur. We developed two methods leveraging care site data: ClinicScan identifies where phenotype documentation occurs, and ClinicWAS identifies specialty utilization patterns associated with a risk factor.

**Materials and Methods:** We extracted care sites for each clinical encounter at an academic medical center and mapped each to a clinical specialty. ClinicScan summarizes the specialty distribution of a user-specified diagnosis; ClinicWAS fits a logistic regression for each care site to identify specialty encounters associated with a user-specified risk factor. We applied ClinicScan to depression to test whether requiring a psychiatry encounter strengthened the association between a polygenic risk score (PRS) and a depression phenotype, and ClinicWAS to a coronary heart disease (CHD) PRS to identify sites enriched for high-risk patients.

**Results:** Across 64,983,257 encounters, 2,544 care sites mapped to 57 specialties. Most depression diagnoses occurred in primary care (30.3%) and psychiatry (19.8%). Requiring a psychiatry encounter strengthened the PRS-phenotype association (OR=1.30, 95% CI 1.26–1.35) versus two or more diagnosis codes alone (OR=1.21, 95% CI 1.19–1.24). CHD ClinicWAS identified 19 associated care sites, including 5 catheterization labs. Men and women with high genetic risk (PRS≥95th percentile) underwent catheterization for CHD 3.1 (1.5–4.6) and 4.6 (2.5–6.7) years earlier than normal-risk participants, respectively.

**Discussion:** Care site data capture phenotype heterogeneity that otherwise distorts EHR-based phenotypes and obscures high-risk subpopulations.

**Conclusion:** Clinical care site data are an under-utilized resource in EHR-linked biobanks.

## Background and Significance

Hospital-based biobanks, in which genetic samples are collected from participating patients and integrated with electronic health record (EHR) data from the same individuals, are increasingly represented in genomic research. Compared with traditional case-control recruitment strategies, biobanks confer the advantage of large sample sizes and readily available, longitudinal clinical data that can be leveraged for high-throughput clinical phenotyping.^1,2^ Traditional EHR phenotyping involves identifying affected patients using recorded diagnosis data such as billing codes.^3,4^ The increasing volume of data available in EHR-linked biobanks has enabled the development of even more granular phenotypes such as disease severity and timing.^5–7^ These “deep” phenotypes allow researchers to better characterize clinical heterogeneity within real-world populations of patients affected by a given condition. Care site data capture where and how patients encounter the health system and represent an under-utilized resource for resolving within-phenotype heterogeneity.^2,8^ A patient with hypertension may receive the same diagnosis code during chronic primary care management or an acute emergency presentation, yet the underlying clinical context differs substantially. Linking encounters to their care sites can therefore refine EHR-based phenotypes and surface healthcare utilization patterns that diagnosis codes alone cannot capture. One limitation, however, is that care site names are not mapped to a controlled vocabulary, so the same type of clinic may appear under different labels across health systems. Standardizing care sites by defining the underlying clinical specialty (e.g., cardiology) of each site is one approach to enable cross-system comparisons. In this study, we demonstrate how structured data such as demographics and billing patterns can be leveraged to annotate care sites by specialty, thereby enhancing their utility for phenotyping and enabling cross-system comparisons.

We develop a data-driven method for classifying clinical care sites according to underlying specialty and demonstrate its research utility through three applications: mapping depression-related care, enhancing genetic cohort construction, and developing a clinic-wide association study approach. Our first application focuses on depression, a heterogenous condition that is often underrecognized and misdiagnosed. Using a curated map linking clinical care sites to specific clinical specialties, we identify clinical settings enriched for depression diagnoses and antidepressant prescriptions (ClinicScan). Next, we demonstrate that incorporating information on psychiatric clinical care settings enables the construction of a depression cohort that is more enriched for depression polygenic risk than one defined using diagnostic codes alone. Finally, we test a clinic-wide association study (ClinicWAS) approach to identify associations between genetic risk for coronary heart disease (CHD) and specialty encounters. To enhance portability of this approach across biobanks, we provide a resource that links commonly documented billing codes to clinical care settings. Our findings underscore the value of care site data in improving the precision and impact of EHR-based genetic studies.

## Materials and Methods

### Clinical data

This study used deidentified EHRs from Vanderbilt University Medical Center (VUMC), a large academic medical center in Nashville, Tennessee. Clinical data were extracted from the Synthetic Derivative (SD) using the Observational Medical Outcomes Partnership (OMOP) Common Data Model (CDM).^9,10^ The SD is a de-identified copy of the VUMC EHR, which includes structured data and unstructured data. EHRs from a total of 3,213,825 individuals were included, spanning from 1998 to 2023.

The key resource for this study was the “care site” OMOP table, which lists physical and organizational settings where clinical care is administered throughout a health system, such as hospitals, wards, and clinics.^11^ The OMOP “visit occurrence” table contains records of clinical encounters, which can be linked to care sites using a unique encounter-specific identifier. A total of 2,544 care sites were manually mapped to 57 clinical specialties by medical students AML and JPS. Specialty labels were based primarily upon manual review of descriptive information available for each care site. In cases in which the specialty was not obvious based on these fields, the label was inferred after review of data commonly documented at that site such as patient demographics, care setting, and billing codes. Care sites associated with multiple specialties were labeled accordingly (e.g., the “VANDY ALLERGY SINUS” care site assigned labels for both allergy/immunology and pulmonology). Further details on the mapping process can be found in **Supplemental Table 1.** Encounter records with no specified care site, as well as those unmappable to a clinical specialty, were excluded from downstream analyses.

For each care site, the most commonly billed diagnosis and procedure codes were identified by mapping encounter records to Current Procedural Terminology (CPT), version 4, and International Classification of Diseases (ICD), versions 9 and 10, codes using the visit_occurrence_id OMOP field. ICD codes were mapped to “phecodes”—groups of related codes representing clinical phenotypes—using the PheWAS R package.^12,13^ Encounters were labeled as inpatient, outpatient, emergency, or unspecified using the visit_type_concept_id OMOP field. Care sites were categorized as sex-specific (male or female), age-specific (adult or pediatric), or setting-specific (outpatient, inpatient, or emergency) if any category exceeded 95% of all clinical encounters.

### Genetic data

Genomic data for this study were obtained from BioVU, the biorepository at VUMC that links leftover blood samples obtained during routine clinical care with de-identified EHRs from the SD.^14^ Genotyping was performed using the Illumina Multi-Ethnic Genotyping Array (MEGA^EX^) and quality controlled using standard procedures as previously described.^15^ Genetic ancestry determination was conducted using principal component analysis (PCA) in FlashPCA 2.0.^16^ Ancestry cluster boundaries were based on proportions from the center of mass of one reference population to another, as previously described.^15^ Imputation was performed using the Michigan Imputation Server^17^ using the Haplotype Reference Consortium reference panel, and related individuals were removed using an identity by descent (IBD) proportion filter of 0.2. A total of 66,917 unrelated individuals with genetic similarity to the 1000 Genomes “Utah residents with Northern and Western European ancestry” (CEU) reference panel^18^ were retained for analysis. After restricting to bi-allelic SNPs and removing SNPs significantly associated with genotyping batch, 6,360,678 SNPs with minor allele frequency (MAF) >0.005 remained.

### Polygenic risk scores

Polygenic risk scores (PRS) for depression and coronary heart disease (CHD) were generated using PRS-CS with default parameters.^19^ For the depression PRS, publicly available summary statistics from a genome-wide association study (GWAS) of over 1.3 million European-ancestry participants were retrieved (371,184 with depression).^20^ This GWAS included a total of 7,547,115 common (minor allele frequency >1%) variants, which were intersected with the quality-controlled BioVU genotyping data. There was a total of 785,280 variants included in the PRS. For the CHD PRS, summary statistics were retrieved for a GWAS of over 1.2 million European-ancestry participants (181,522 CHD cases).^21^ Of 8,365,359 variants in the GWAS, there were 781,117 variants after intersecting with the quality-controlled BioVU set.

### ClinicScan of depression care

Care sites associated with depression diagnoses and antidepressant medication records were identified by mapping each source of clinical data to care sites and specialties. For the depression diagnosis analysis, depression ICD-9 and ICD-10 billing codes mapping to the depression phecode (296.2) were extracted.^13^ Then, each patient’s earliest depression ICD-9 or ICD-10 code was identified and linked to clinical encounters occurring on the same date as the diagnosis (to capture more data than available when linking by the unique visit occurrence identifier). Clinical encounters mapping to “Unclear” or “Administrative” specialties were excluded. Next, for each care site, the proportion of patients first receiving a diagnosis code in that site was calculated and normalized by (divided by) the EHR-wide number of patients with a clinical encounter at that site.

For the antidepressant analysis, antidepressant medication records were extracted using regular expression matches to commonly prescribed antidepressants. Medication records classified as “Prescription written,” “Physician administered drug (identified as procedure),” “Prescription dispensed in pharmacy,” and “Inpatient administration” were retained for analysis. The same mapping approach as described for diagnoses was then applied for patients receiving antidepressant prescriptions.

### Comparing depression phenotype definitions

Pairwise comparisons of PRS across three depression phenotype definitions were conducted using linear regression with covariates for EHR-recorded sex, median age at encounter, and the top ten genotyping principal components. A total of five depression phenotype definitions were separately assessed for association with depression PRS using logistic regression adjusting for the same covariates. Odds ratios were compared to determine the relative strength of PRS-depression associations across phenotype definitions. Individuals with only a single depression code were excluded from PRS analyses.

### ClinicWAS of coronary heart disease (CHD)

A clinic-wide association study (ClinicWAS) fits a logistic regression for each site with ever having an encounter at a specified care site as the dependent variable and a user-specified independent variable, such as a risk factor. Controls were defined as patients with clinical encounters falling within both the active date range of each care site and the 1st to 99th percentile age range of that care site’s patient population, providing approximate age-matching. For care sites where >95% of patients were male or female, participants of the other sex are excluded. The PheWAS R package was adapted to perform association analyses between each exposure and a care site.^12^

We performed ClinicWAS in adult (>18 years old) participants to assess the association between elevated genetic risk for CHD and care site encounters. Care sites were excluded from analysis if they had fewer than 100 unique participants. The independent variable was a binary variable defined as “high risk” if the PRS value for either condition was above the 95^th^ percentile of the distribution. This risk threshold has been employed by a clinical implementation study led by the Electronic Medical Records and Genomics (eMERGE) Network.^22^

The association between CHD genetic risk on age at first cardiac catheterization was assessed by restricting analyses to a subset of patients with encounters at care sites labelled as interventional cardiology or cardiac catheterization care sites. Age was defined as the first instance of an encounter on the same day as an ICD code for coronary heart disease which included phecodes for: unstable angina (411.1), myocardial infarction (411.2), angina pectoris (411.3), coronary atherosclerosis (411.4). All analyses were adjusted for EHR-recorded sex, individual median age at clinical encounter, and the top ten principal components.

### Identifying patterns of CPT billing codes across specialties

To support cross-institutional portability, we identified specialty-specific CPT signatures. For each care site, we determined the number of unique CPT codes accounting for 99% of billing activity. We then computed term frequency (TF) and inverse document frequency (IDF) for each CPT code by specialty, borrowing measures traditionally used for natural language processing, to identify billing patterns most specific to a given specialty.^23^ TF was the proportion of billings of a given code within a specialty. IDF was the log of the total number of specialties divided by the number of specialties in which the code appeared. The TF-IDF product captures specialty-specificity of a CPT code.

### Statistical analysis software

All analysis was performed using R versions 4.4.0 and 4.4.2.^24^

### IRB information

This study was approved by the Vanderbilt Institutional Review Board (IRB) and designated “not human subjects” research under protocol numbers 190418 and 160650. All research procedures were conducted in accordance with the ethical guidelines outlined by the IRB.

## Results

### Clinics are diverse and show the variation of patient encounters within EHR-based biobanks

In the VUMC EHR, there were 64,983,257 clinical encounters for 3,213,825 patients between 1998 and 2023 (**Figure 1**). The care site of the encounter was documented in 79.7% of encounters. Of the 2,544 unique care sites, 2,189 (86.0%) sites could be mapped to one of 57 underlying specialties. Excluded sites included 321 that were not specific to any given specialty and 34 that were administrative and primarily involved with care coordination activities (e.g., scheduling, discharge) rather than patient care. Most care sites were instituted after VUMC’s implementation of the Epic EHR system in 2017 (**Supplemental Figure 1**).^25^

**Figure 1:**
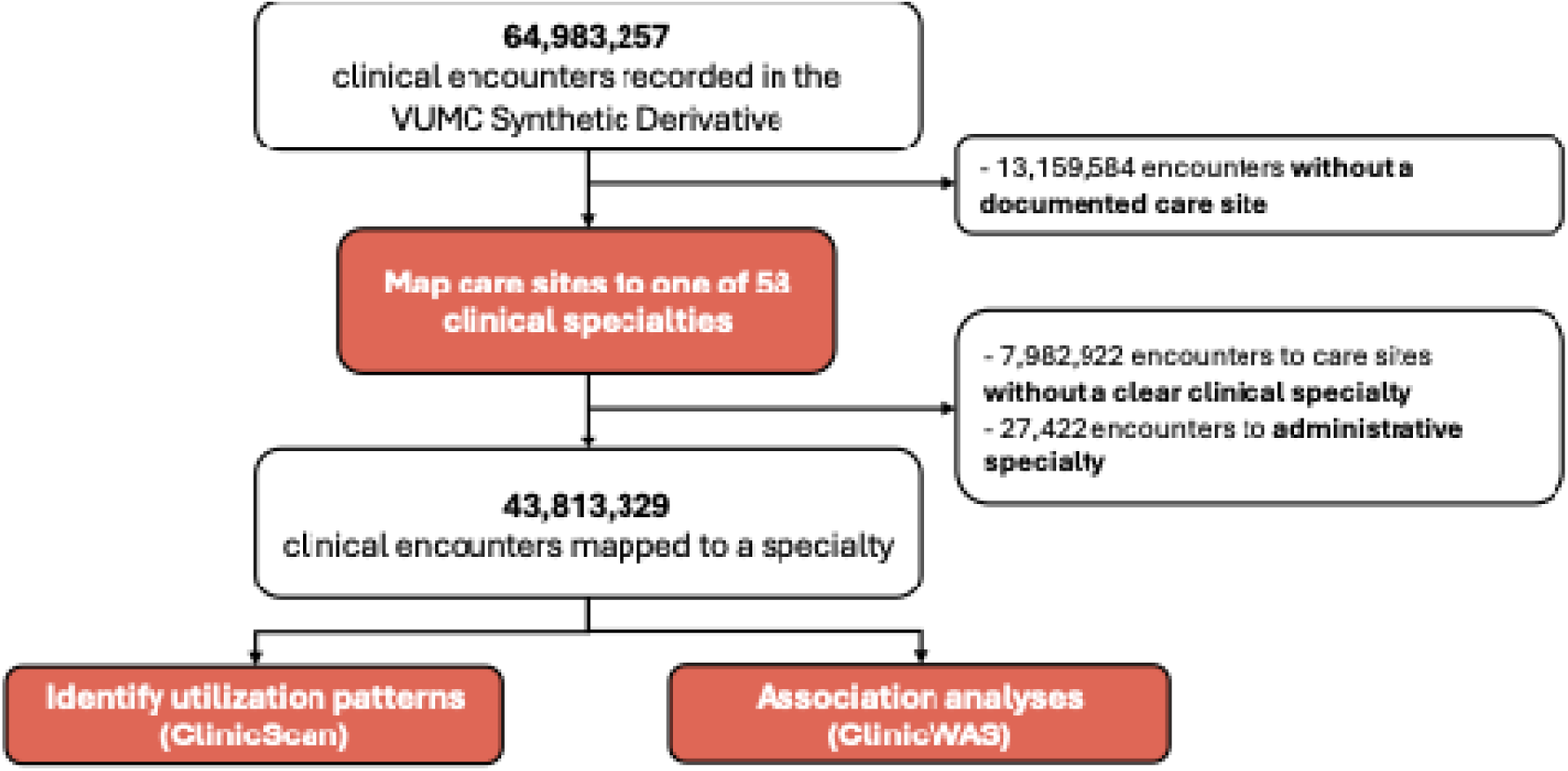
Study overview.

After restricting to encounters mapped to non-administrative clinical specialties, there were 43,813,329 encounters and 2,959,903 patients remaining. The majority of encounters were for adult patients, female patients, and were conducted in the outpatient setting (**Supplemental Table 2**). Care sites had a median of 2,725 (IQR, 281 – 15,361) encounters for 1,195 (174 – 6,379) unique patients. Care sites were primarily age-specific (59.7% adult-predominant sites, 16.7% pediatric-predominant sites) but were not frequently gender-specific (8.7% female-predominant sites, 2.0% male-predominant sites). Across care sites, circulatory diagnoses predominated, with 35.2% of care sites having this as the most common diagnostic category (**Supplemental Table 2**).

The most frequently visited specialties were primary care (20.0% of all encounters), cardiology (7.5%), radiology (6.9%), hematology/oncology (6.9%), and orthopedic surgery (6.4%). Across the top 25 specialties, the majority of clinical encounters were outpatient with the exception of Emergency Medicine and General Surgery, which had an enrichment of emergency and inpatient encounters (**Figure 2**). Care sites were predominantly mapped to a single primary specialty, but 322 (14.7%) sites were mapped to multiple specialties (**Supplemental Figure 2)**. The most common combination of specialties was primary care and urgent care, with 48 sites mapping to both specialties. Among the top 50 most visited sites for each specialty group (Medical, Surgical, or Other), the diagnosis category of the most common ICD code corresponded well to the specialty annotation (**Figure 3**). Full descriptive statistics for each care site and specialty can be found in **Supplemental Data 1**.

**Figure 2:**
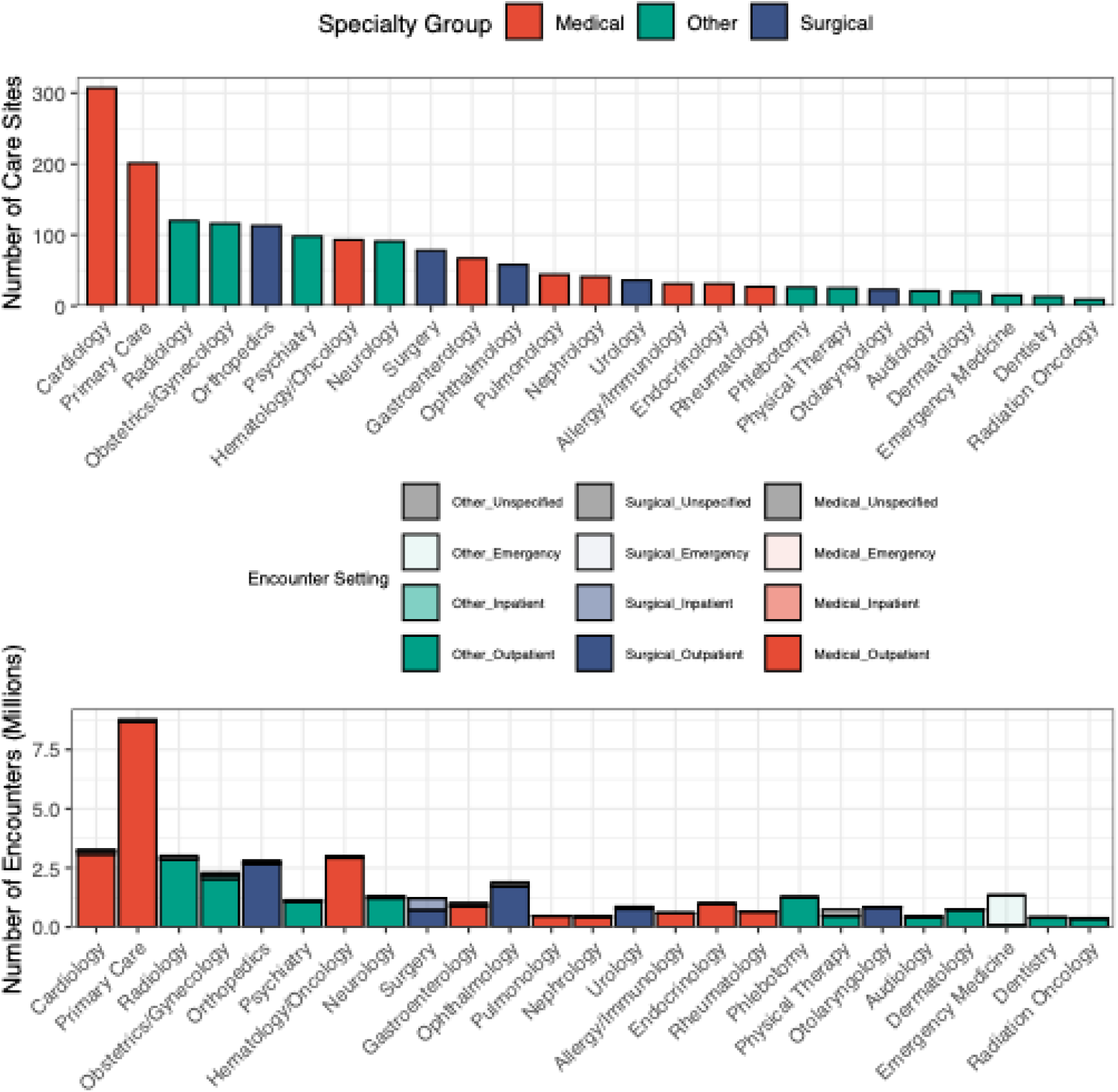
Care site specialty map enables characterization of the clinical contexts from which electronic health record (EHR) data are derived. **Top**: Number of distinct care sites identified per specialty, color-coded by encounter setting (outpatient, inpatient, emergency, or unspecified). **Bottom**: Number of clinical encounters to each clinical specialty, summed across all care sites mapped to that specialty.

**Figure 3:**
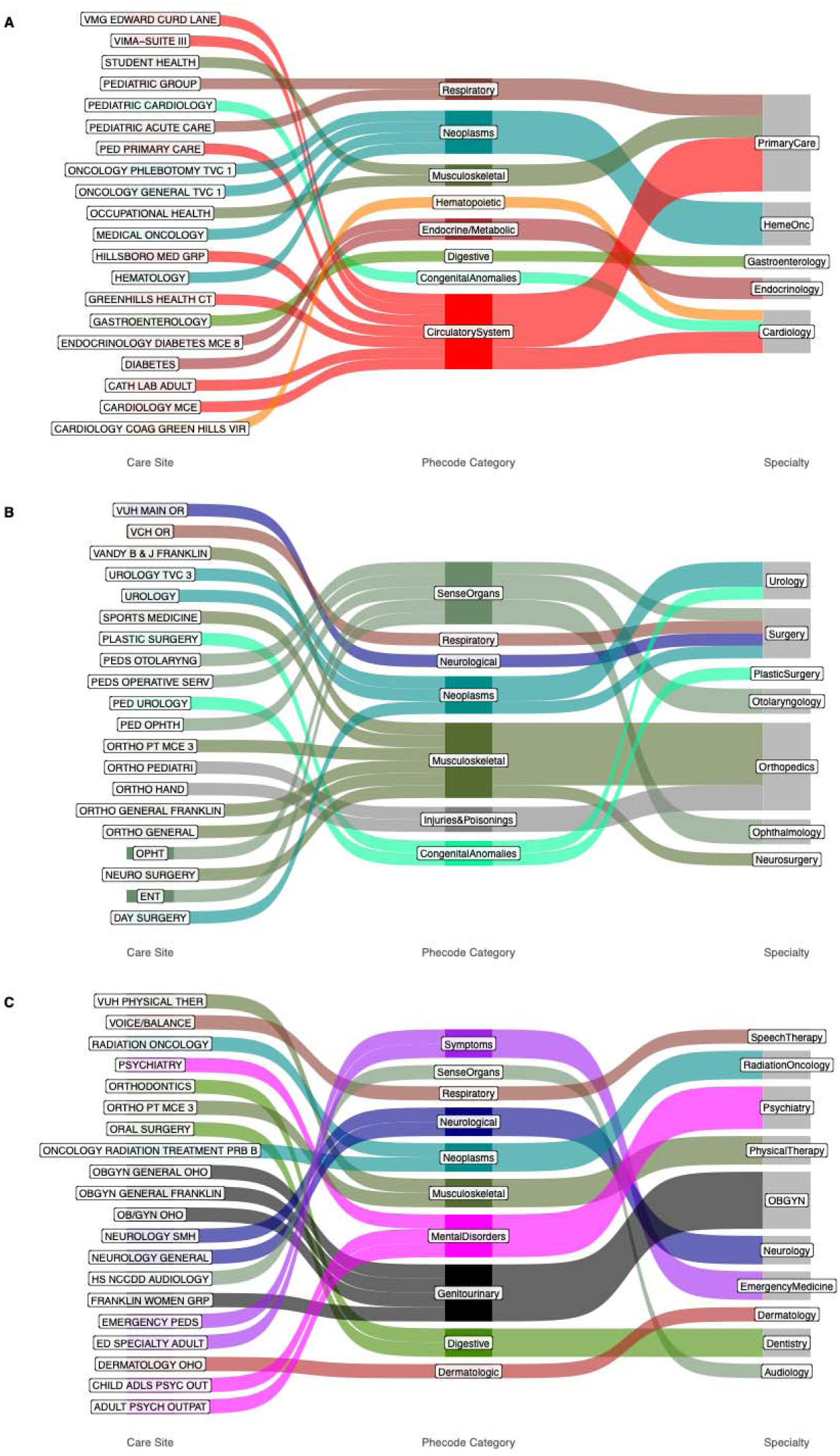
Care site specialty mapping reflects expected relationships between specialties and diagnoses. The top 20 most commonly visited care sites (leftmost nodes) associated with **A)** medical**, B)** surgical, and **C)** other specialties are shown, along with links to the most commonly documented diagnostic categories based on Phecode classifications per care site (center nodes) and links between care sites and mapped clinical specialties (rightmost nodes).

### Mapping clinical care sites informs EHR-based phenotyping of depression

Depression is a heterogeneous condition with substantial genetic and phenotypic overlap with other psychiatric conditions.^26–28^ While the gold standard is a DSM-compliant diagnosis made in a mental health setting, depression is commonly diagnosed in primary care, and only 21% of antidepressant prescriptions are written by mental health providers.^29^ We therefore hypothesized that most depression diagnoses and prescriptions in the EHR would be documented outside of psychiatric settings.

We evaluated this hypothesis using a clinic-wide scan (“ClinicScan”) to determine the proportion of patients’ earliest EHR depression diagnoses and antidepressant prescriptions documented in each clinical care site. Across 149,607 individuals with at least one depression diagnosis code **(Supplemental Table 4),** we found that substantial proportions of these diagnoses were documented in primary care (30.3% of diagnoses), emergency medicine (10.6%), and surgery (9.2%) settings, in addition to psychiatry settings (19.8%, **Figure 4**). When normalizing by per-site and per-specialty individual count to account for the non-uniform distribution of clinical encounters across care sites **(Figure 2)**, we observed an attenuation of the relative overrepresentation of these non-psychiatry specialties, an increase in the relative enrichment in psychiatry settings (most highly *enriched* specialty, 17.4% of diagnoses per 100,000 patients specialty-wide), as well as the emergence of geriatric medicine as the second most highly enriched specialty (8.3% of diagnoses per 100,000 patients specialty-wide, **Figure 4**). We observed a similar pattern in a ClinicScan of 215,955 individuals with at least one antidepressant prescription (**Supplemental Table 4, Supplemental Table 5**), with the largest proportions of medications prescribed in primary care (27.6% of prescriptions), neurology (13.7%), psychiatry (11.2%), and surgery (8.0%) settings and increased enrichment in psychiatry settings when accounting for overall specialty encounter frequencies (9.8% of prescriptions per 100,000 patients specialty-wide, **Supplemental Figure 3**).

**Figure 4:**
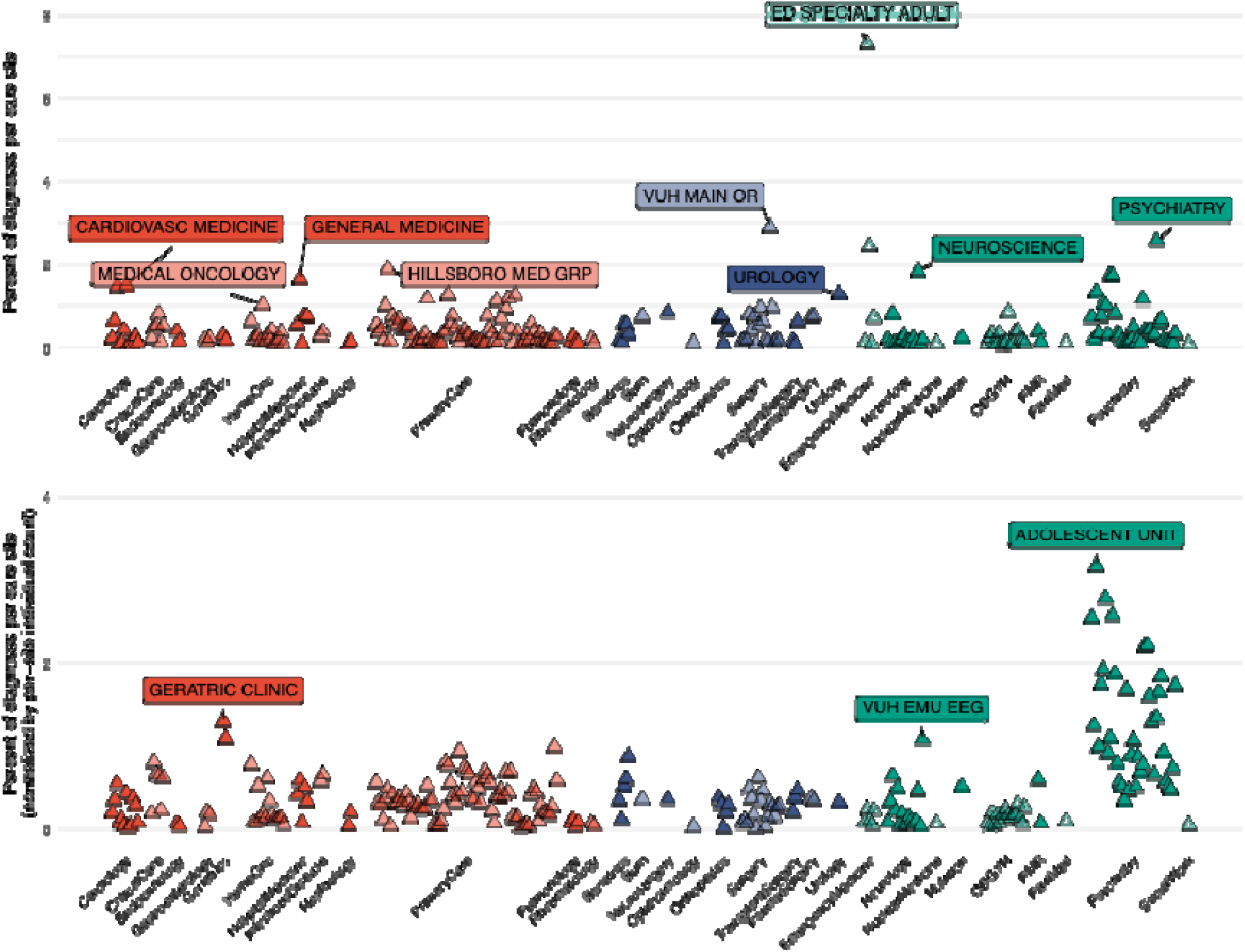
Depression diagnosis ClinicScan plot showing variation in depression diagnoses across the health system. Each point represents the percentage (upper panel) or normalized percentage (lower panel) of depression diagnoses documented in that particular care site. Only the earliest documented diagnosis for each patient was considered. Clinical specialties are grouped into medical (red), surgical (blue), and other (green) specialties. Only sites with at least 100 diagnoses are included, and the top site per specialty with a percentage of ≥1 is labeled.

Next, we evaluated whether mapping clinical care sites to specialties would identify individuals with a higher burden of depression-associated genetic risk, as quantitated by a depression PRS. We examined associations between depression PRS and five depression phenotype definitions in a cohort of 61,511 genotyped individuals, of whom 10,414 had at least two depression diagnosis codes on separate days (**Supplemental Table 4**). We first considered two definitions of depression: one requiring at least two depression ICD-9 or ICD-10 codes and at least one psychiatry clinical encounter, and one only requiring two codes. Both depression definitions had significantly greater mean standardized depression PRS than individuals without a code when adjusting for sex, median age at encounter, and the top ten genotyping principal components (p<1e-5, **Figure 5A**). Furthermore, among patients with at least two depression codes, those who had evidence of a psychiatry encounter had greater depression average PRS values than those who did not (beta = 0.10, 95% CI = 0.06-0.14, **Figure 5A**). Next, we constructed additional depression definitions, requiring two, three, four, or ten depression codes, respectively. As expected, requiring only two depression codes resulted in the smallest magnitude of association between depression PRS and phenotype (odds ratio per SD unit increase in PRS (OR) = 1.21, 95% CI = 1.19-1.24, **Figure 5B**). Requiring an increasing number of depression codes led to larger magnitudes of association between PRS and diagnosis. However, the strongest association between depression PRS and depression phenotype was observed when requiring a psychiatry clinical encounter in the depression case definition (OR =1.30, 95% CI = 1.26-1.35).

**Figure 5:**
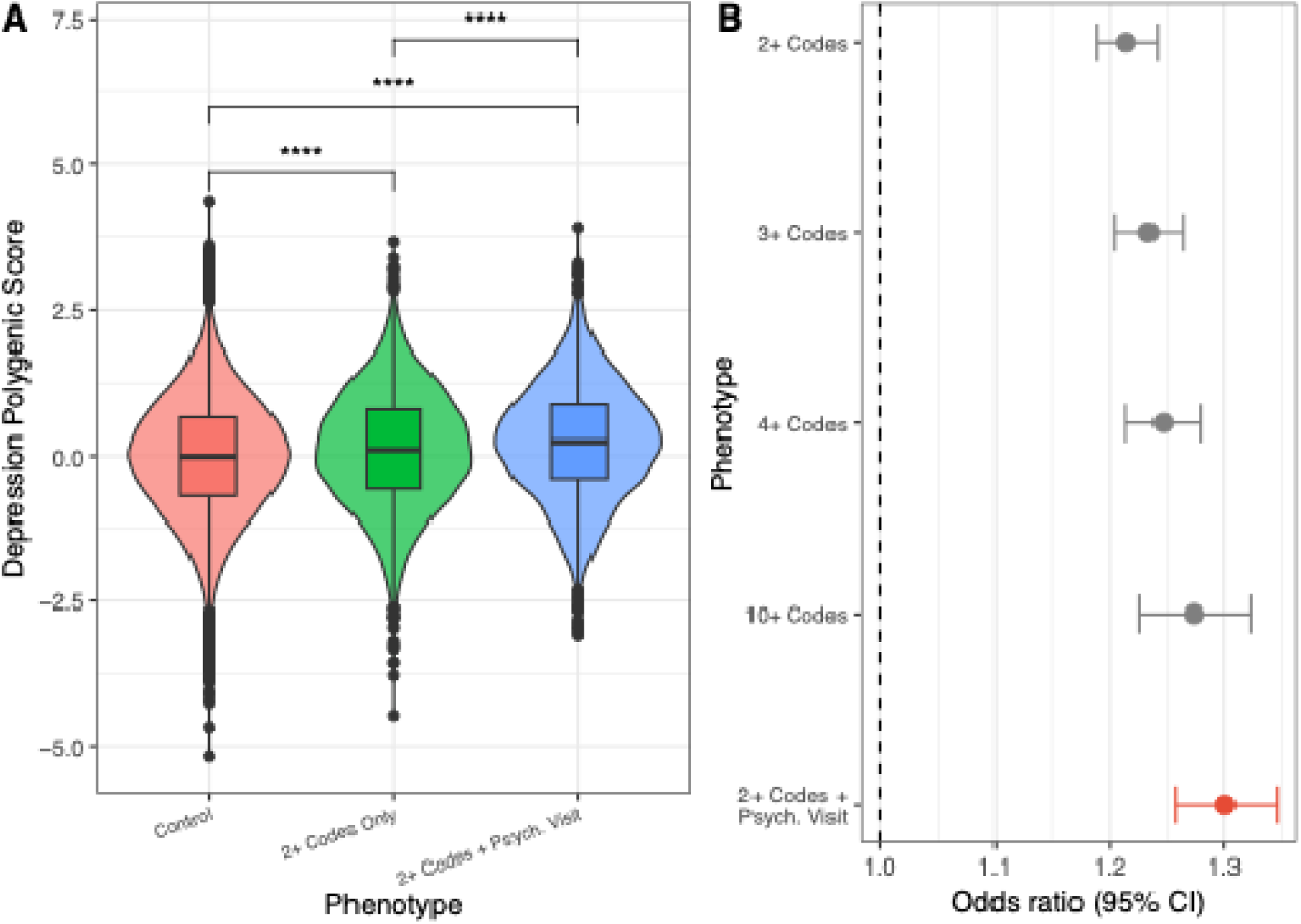
Incorporating psychiatric care information increases magnitude of associations between depression polygenic risk scores and depression phenotype. **A)** Violin and box plots of standardized depression polygenic risk scores (PRS) across two depression cohorts (2+ Codes + Psych Visit, N=3,690, 2+ Codes Only, N=6,724) and individuals with no depression ICD code (Controls, N=51,137). Asterisks indicate significant difference in PRS between groups, after adjusting for EHR-recorded sex, individual median age at encounter, and top ten genotyping principal components. **B)** Forest plots illustrating logistic regression of varying depression phenotype definitions as the dependent variable and standardized polygenic risk scores as the independent variable, with the same covariates as used in panel **A**.

### ClinicWAS of genetic risk for heart disease identifies acute and post-acute cardiac care sites for sub-population analysis

We next applied ClinicWAS to characterize healthcare utilization associated with polygenic risk for coronary heart disease (CHD).^30^ Among adult BioVU participants (N=55,020), participants at highest polygenic risk for CHD (PRS > 95^th^ percentile) were more likely to have encounters at 19 clinical care sites at the Bonferroni-corrected significance threshold (p<3.2×10^−5^). These sites were primarily cardiology and vascular surgery specialty locations (**Figure 6A and 6B, Supplemental Table 6**). Associated care sites included those focusing on acute management (e.g., cardiac catheterization lab) and post-event care (e.g., cardiac rehabilitation clinic), thereby demonstrating the utility of ClinicWAS for identifying more granular sub-populations for further study. To demonstrate this use case, we identified the subset of participants who had an encounter for CHD at one of 13 cardiac catheterization sites and tested how genetic risk was associated with age at catheterization (**Supplemental Table 7)**. In the population undergoing catheterization (N=5,120), high-risk men and women had their first event at our institution a mean of 3.1 (1.5 - 4.6) and 4.6 (2.5 - 6.7) years earlier, respectively, than lower risk participants (**Figure 6C**). Similarly, high polygenic risk participants were more likely to meet criteria for early-onset CHD (OR = 2.06, 95% CI = 1.59 - 2.67). Together, these results show how discovery-oriented testing across care sites can characterize the clinical contexts in which underlying genetic risk manifests and identify sub-populations for further evaluation.

**Figure 6.**
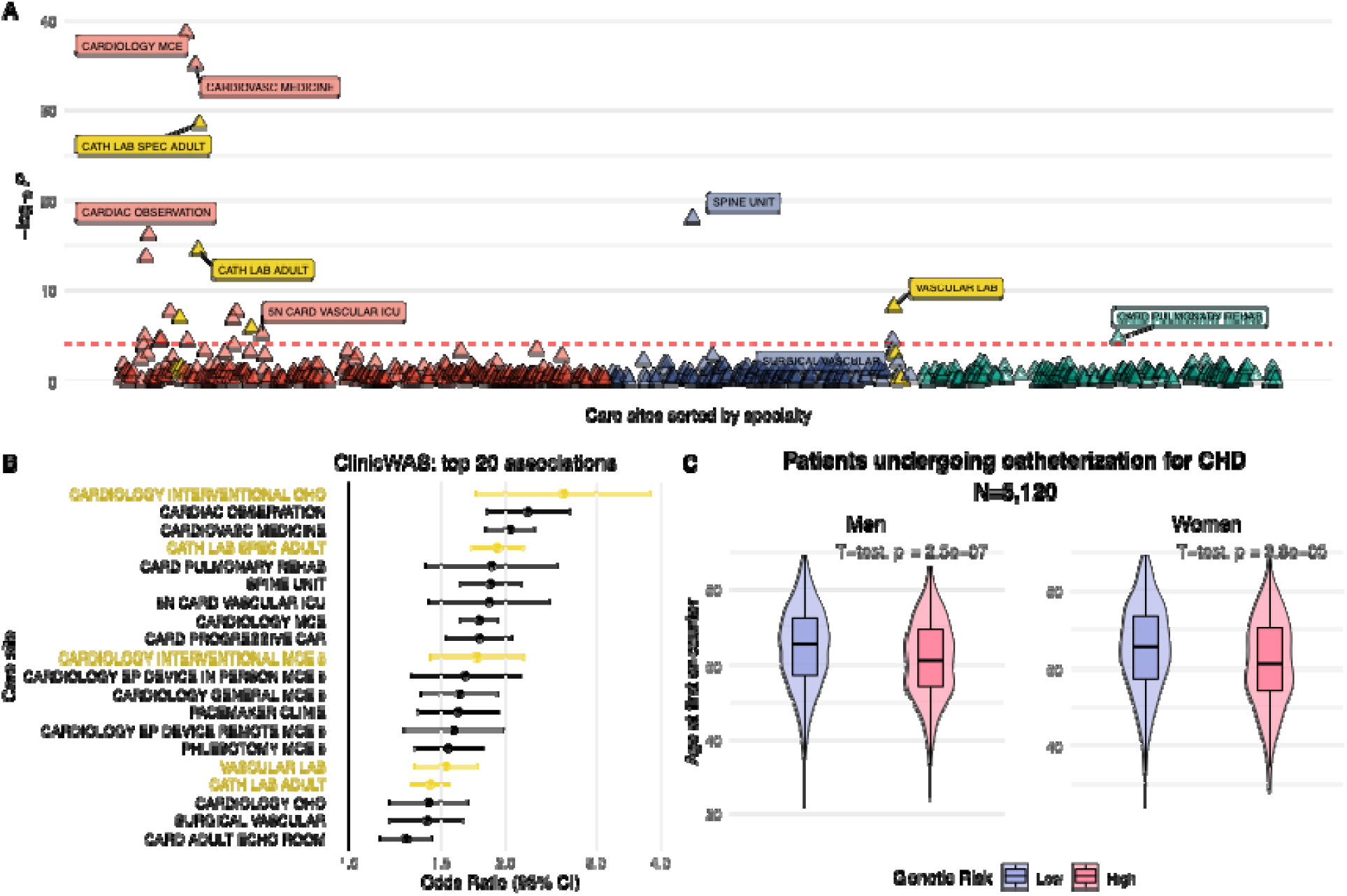
Genetic risk for coronary heart disease (CHD) is associated with cardiac catheterization and early-onset CHD among patients undergoing catheterization at VUMC. **A)** Manhattan plot of the ClinicWAS of genetic risk (PRS_CHD_ > 95^th^ percentile) in the adult cohort (N=55,020). The x-axis represents the care sites, sorted by annotated specialty, while the y-axis shows the statistical significance of the association between genetic risk (>95^th^ percentile) and each care site. Cardiac catheterization sites are highlighted in yellow, while other points are colored according to specialty group: medical, surgical, or other. The horizontal line indicates the Bonferroni-corrected significance threshold. **B)** Forest plot showing the top 20 associations between genetic risk and care sites, sorted by odds ratio. The y-axis is the name of the care site. **C)** Violin plots show the age distribution of high- and low-risk patients at their first encounter at an interventional cardiology or catheterization lab care site for CHD (N=5,120). Results are sex-stratified.

### Specialties carry CPT signatures that can be used to enhance portability

We hypothesized that specialty sites carried distinct billing signatures that could be used to classify care sites according to specialty at other institutions. A median 775 (371 - 1,796) unique CPT codes were billed for each care site. Within each site, however, billing was concentrated with a median 28 codes (9 - 79) accounting for 99% of that site’s billing volume (**Supplemental Figure 4**). Term frequency (TF) and inverse document frequency (IDF) were calculated for each unique CPT code to capture the distinctiveness of CPT codes for each specialty (**Supplemental Data 2**). We identified CPT codes that were billed most frequently (i.e., highest TF) within each specialty (**Supplemental Figure 5**). After IDF-weighting, specialties service distinct patient populations had higher peak TF-IDF codes, indicating more specialty-specific billing patterns (**Supplemental Figure 6**). The median TF-IDF score across all specialty-CPT code pairs was 5.4×10^−4^. The most specialty-specific CPT codes had TF-IDF scores of 0.32 for nursing home care (99310) in geriatrics and 0.10 for prenatal visits (0502F) in obstetrics and gynecology (**Supplemental Figure 6**). These specialty-level CPT signatures can be used by researchers at other institutions to infer the underlying clinical specialty of unannotated care sites based on their billing patterns, enabling cross-system applications of the methods we describe.

## Discussion

In this study, we provide a comprehensive characterization of the clinical care sites in one of the largest hospital-based, EHR-linked biobanks. We then develop two methods: ClinicScan, a descriptive approach to summarize clinical settings administering specific diagnoses or treatments, and ClinicWAS, an analytical approach to identify clinical settings enriched for genetic risk for a particular condition.

First, using depression as a use case, we illustrate how knowledge of clinical care sites and specialties administering depression care can be used to gain insights into patterns of depression diagnosis and treatment throughout the health system. We then demonstrate how this information strengthens the association between EHR-based depression phenotypes and depression polygenic risk, consistent with improved concordance with the case definitions used in depression GWAS. Given the complexity of depression and its diagnosis, a research-quality depression phenotype would ideally involve a diagnosis in a psychiatry setting using gold-standard diagnostic criteria. We show that by including evidence of psychiatric specialty care in the phenotyping criteria, the resulting depression cohort is more highly enriched for depression polygenic risk, compared with phenotyping definitions using billing codes only. This finding held true even when requiring up to ten depression codes on separate days. These findings exemplify how the specialty-level information provided through our clinical care site mapping procedure can enhance the rigor of EHR-based phenotyping approaches for genetic research.

Second, we use ClinicWAS to characterize the clinical contexts in which polygenic risk for a given condition manifests as healthcare utilization. Using the same design as the PheWAS approach, ClinicWAS identifies care sites where high-risk patients are more likely to encounter the health system, which can in turn be used to specify more granular phenotypes. Applying this to CHD polygenic risk, we identified care sites corresponding to distinct stages and severity of disease, including acute presentation (i.e., catheterization sites) and post-event management (e.g., cardiac rehabilitation) which could enable the construction of stage-specific phenotypes rather than a single binary case definition. Among catheterized patients, those with greater genetic liability to CHD had events at significantly younger ages than lower-risk patients. These differences are conditional on having presented for catheterization and characterize the timing of clinical presentation among cases rather than disease incidence in the general population. Whether prospective polygenic screening would shift these trajectories requires interventional study, such as the ongoing eMERGE PRS implementation trial.^31^

Because EHR-linked biobanks represent real-world clinical populations, these approaches can identify care settings that more commonly encounter patients with a given trait. By identifying these settings, ClinicScan and ClinicWAS can enable more targeted recruitment for pragmatic clinical trials which are conducted during the process of routine care.^32^ Tertiary care centers provide an overwhelming variety of clinical settings to recruit from but investigators must refine their recruitment site selection for feasibility.^33^ The approaches outlined in this study can be used by study investigators in the preparation phase to identify care sites with adequate numbers of the target patient population.^34^ While we utilized genetic risk as the test variable for ClinicWAS, researchers could implement a ClinicWAS for predicted eligibility status, defined using study inclusion and exclusion criteria, to identify care sites most likely to care for the target population. In summary, utilizing care site data can optimize recruitment strategies by identifying care sites where eligible participants are more likely to be found, increasing the efficiency and cost-effectiveness of pragmatic trials.

Our study has several limitations. First, in our database of clinical encounters, 22.2% of clinical encounters had no specified care site. Furthermore, of the 2,544 unique care sites identified, 14% could not be mapped to an underlying specialty due to unclear or nonspecific care site names (e.g., “No Location,” “Unspecified Clinic,” “O/P Vandy Default,” “MCE 3”). While missing data is an inherent limitation of all EHR-based studies, we believe the available data are sufficient to broadly illustrate patterns of specialty-specific care across the health system. Second, care sites are administrative constructs and may not capture granularity or specificity comparably across institutions. Thus, our manually curated map of care sites to clinical specialties is specific to the VUMC system. However, we provide the list of procedure codes commonly documented in each mapped specialty **(Supplemental Data 2)**, along with indices of their frequency and specificity to clinical specialties. These data can be utilized by researchers at other institutions, as CPT codes are portable across EHRs. Additional work is needed to determine how well these CPT codes capture clinical specialties in independent EHR systems.

## Conclusion

Taken together, our findings demonstrate the importance of incorporating clinical settings when defining and refining phenotypes in EHR-based biobanks. We demonstrate that this information can be used to describe patterns of clinical care, inform EHR-based phenotyping, and identify novel associations between genetic risk and specialty care. These patterns can be used in a variety of precision medicine applications including enhancing recruitment for pragmatic trials and mitigating setting-specific ascertainment bias in biobank-based studies.

## Supporting information

Supplemental Data 1. Source data for care sites and associated descriptive statistics

Supplemental Data 2. Frequency, TF, IDF, and TF-IDF of all CPT codes by specialty

## Acknowledgements

**General**: Vanderbilt University Medical Center BioVU projects are supported by numerous sources, such as institutional funding, private agencies, and federal grants. These sources include Shared Instrumentation Grants S10OD017985, S10RR025141, and S10OD025092, as well as Clinical and Translational Science Awards UL1TR002243, UL1TR000445, and UL1RR024975, from the NIH. Genomic data are also supported by investigator-led projects that include NIH grants U01HG004798, R01NS032830, RC2GM092618, P50GM115305, U01HG006378, U19HL065962, and R01HD07471, as well as additional funding sources listed at https://victr.vumc.org/biovu-funding.

**JPS:** R01GM130791, T32GM152284

**AML:** F30MH132165, T32GM152284

**JMS:** None

**TEU:** None

**JFP:** None

**LKD:** R01MH137220

**JDM:** Institutional support from UTSW

## Author contributions

Concept and design: Shelley, Lake

Acquisition, analysis, or interpretation of data: All authors

Drafting of the manuscript: Shelley, Lake

Critical revision of the manuscript for important intellectual content: All authors

Statistical analysis: Shelley, Lake

Administrative, technical, or material support: Mosley, Davis, Sealock

Supervision: Mosley, Davis

## Competing interests

**JPS:** None

**AML:** None

**JMS:** None

**TEU:** None

**LKD:** None

**JFP:** Myome, Inc (consulting)

**JDM:** None

## Data Availability Statement

Data are available from Vanderbilt University Medical Center with institutional restrictions that apply to the acquisition, use, and dissemination of data. To request reasonable access to data for work conducted in a non-profit academic setting, please contact the Vanderbilt Institute for Clinical and Translational Research and request an application to the Integrated Data Access and Services Core. All code used for analysis can be found at https://github.com/amlake/care_sites.

## Notes

### Competing Interest Statement

Dr. Josh Peterson has provided consulting for Myome, Inc.

### Author Declarations

This study was approved by the Vanderbilt University Institutional Review Board (IRB) and designated as not human subjects research under protocol numbers 190418 and 160650. All research procedures were conducted in accordance with the ethical guidelines outlined by the IRB.

