## Supplemental Information for "Clinical care site data integration reveals heterogeneity in EHR phenotyping and healthcare utilization patterns"

### Supplemental Table 1: Description of each assigned care site specialty category, including keywords and rules used to label each specialty.

| **Specialty** | **Keywords** (case insensitive) | **Notes** |
| --- | --- | --- |
| Administrative | scheduling, admitting, registration/regis*, transport |  |
| AllergyImmunology | VASAP, allergy | VASAP = Vanderbilt Asthma, Sinus, and Allergy Program |
| Anesthesiology | Anesthesia/anes, periop, HIRISE, VPEC | HIRISE = High-Risk Surgical Encounter Clinic  VPEC = Vanderbilt Pre-Operative Evaluation Center |
| Audiology | Audiology, cochlear, hear aids |  |
| Bariatrics | Weight loss, bariatric |  |
| Burn | Burn |  |
| Cardiology | Cardiology, Heart, EKG, Holter, event monitor, vascular, cath, cardiovasc, treadmill | All EKG sites are included. Sites with “vascular” include both cardiology and vascular surgery. Unless location_id=Cardiology, label as vascular surgery. |
| CardiothoracicSurgery | Thoracic, cardiac, cardiothoracic |  |
| ColorectalSurgery | Colorectal |  |
| CriticalCare | Critical, ICU |  |
| Dentistry/OMFS | Oral & maxillofacial, orthodontics, dentistry, surgery oral |  |
| Dermatology | dermatology, dermatolog, dermatopath, skin, phottherapy |  |
| DevelopmentalPediatrics | Developmental med/developmental medicine, motor impairment, kennedy, down syndrome, fragile x, prader willi, angelman |  |
| EmergencyMedicine | EM, ED, emergency, emer, lifeflight/life flight |  |
| Endocrinology | Endocrinology/endocrinol, diabetes |  |
| ENT | Otolaryngology, ENT, Craniofacial |  |
| Gastroenterology | Gastroenterology, endoscopy, liver, EGID, IBD, hepatology |  |
| Gender-affirming care | VIVID health Bellevue, transgender |  |
| Genetics | Genetics, genomics |  |
| Geriatrics | Geriatrics, nursing home |  |
| HemeOnc | Hematology, oncology/onc, cancer, chemo, breast, myelo/myelosuppression/meylo, stem cell, sickle cell, CHIIP | Surgical oncology sites are labeled as both Surgery and HemeOnc. |
| HospitalMedicine | Medicine / surgery, med/surg, medicine, general med |  |
| InfectiousDisease | Infectious, infection, infec dise, |  |
| InterventionalRadiology | Interventional radiology |  |
| Neonatology | Nursery/nurs, neonatal/neonatolog/neonatology |  |
| Nephrology | Renal, dialysis, nephrology |  |
| Neurology | Neurology/neuroscience/neuro, epilepsy, EEG, sleep, spina bifida, ALS, MS |  |
| Neurosurgery | Neurosurgery, spine, brachial plexus |  |
| NuclearMedicine | Nuclear medicine |  |
| Nutrition | Nutrition |  |
| OBGYN | OBGYN/OB, gynecology/gyn, CWI, lactation, women, fetal, labor and delivery, partum, midwifery |  |
| OccupationalTherapy | Occupation, OT |  |
| Ophthalmology | Ophthalmology/opht, cornea |  |
| Orthopedics | Orthopedics/orthopaedics, sports, B & J/VB&J |  |
| PainMed | Interventional, pain |  |
| PalliativeCare | Palliative |  |
| Pathology | Pathology/path, lab, cytopath/cyto |  |
| Pharmacy | Pharmacy |  |
| Phlebotomy | Phlebotomy | In addition to primary phlebotomy sites, other phlebotomy sites that appear to be associated with a particular specialty are labeled with both that specialty and Phlebotomy. |
| PhysicalTherapy | Physical therapy, PT |  |
| PlasticSurgery | Plastics/plastic, cosmetic |  |
| PMR | PM&R, rehab |  |
| PrimaryCare | Walk (walk-in clinics), VIMA (Vanderbilt Internal Medicine Associates), Adolescent (adolescent medicine) | Inpatient services are assigned Medicine rather than PrimaryCare |
| Psychiatry | Psych, Behav, VPH, VITA | VITA = Vanderbilt Institute for Treatment of Addiction  Partial = partial hospitalization |
| Pulmonology | Pulmonary, lung, PFT, cystic fibrosis |  |
| RadiationOncology | Radiation oncology, oncology radiation |  |
| Radiology | X-ray, fluoro, PET, MRI, CT, ultrasound, bone density, mammo | In addition to primary radiology sites, other radiology sites that appear to be associated with a particular specialty are labeled with both that specialty and Radiology. |
| Research | Research, CRC |  |
| Rheumatology | Rheumatology/rheum, arthritis |  |
| SocialWork | Social work, case management, care navigation |  |
| SpeechTherapy | Speech, SLP, voice |  |
| Surgery | General surgery, gen surg, holding, recovery, OR/operating | Sites with discernible surgical subspecialties are labeled as such. All others are given the ‘Surgery’ label. |
| Toxicology | Toxicology, pharmacology |  |
| TransplantSurgery | Transplant | Organ-specific sites get this label in addition to the corresponding medical subspecialty. E.g., a renal transplant site gets labeled as both TransplantSurgery and Nephrology. |
| TraumaSurgery | Trauma |  |
| Urology | Urology/urologic, kidney stone |  |
| VascularSurgery | Vascular |  |
| WoundCare | Wound | Exclude if ostomy |

### Supplemental Table 2: Demographic characteristics of clinical encounters in the Synthetic Derivative (N=2,959,903)

| **Characteristic** | **Synthetic Derivative (N=2,959,903)** |
| --- | --- |
| Number of encounters | 43,813,329 |
| Number of care sites | 2,189 |
| **Care setting** |  |
| Outpatient | 89.5% |
| Inpatient | 4.4% |
| Emergency | 2.9% |
| Unspecified | 3.2% |
| **Age of participant** |  |
| 18 or older (adult) | 78.7% |
| Less than 18 (pediatric) | 21.3% |
| **Sex of participant**^a^ |  |
| Female | 57.0% |
| Male | 43.0% |

^a^ Sex refers to EHR-recorded sex, which may not reflect gender identity or sex assigned at birth.

### Supplemental Table 3: Demographic characteristics of clinical care site populations (N=2,189)

| **Characteristic** | **Clinical Care Sites**  **(N=2,189)** |
| --- | --- |
| Encounters per care site (median [25%-75%]) | 2,725 (281 - 15,361) |
| Unique patients per care site (median [25%-75%]) | 1,195 (174 - 6,379) |
| Age (median [25%-75%]) | 51 years old (16 - 61) |
| **Predominant care setting^a^** |  |
| Outpatient | 66.1% |
| Mixed | 25.4% |
| Inpatient | 8.3% |
| Emergency | 0.3% |
| **Age specificity^a^** |  |
| Adult | 59.7% |
| Pediatric | 16.7% |
| Not specific | 23.7% |
| **Sex specificity^a,b^** |  |
| Female | 8.7% |
| Male | 2.0% |
| Not specific | 89.3% |
| **Diagnostic category of most common ICD code** | |
| Circulatory system | 35.2% |
| Respiratory | 18.2% |
| Neoplasms | 16.0% |
| Musculoskeletal | 15.8% |
| Neurological | 14.8% |

^a^ Care sites are defined as age-specific, sex-specific, or setting-specific if any category exceeds 75% of total encounters.

^b^ Sex refers to EHR-recorded sex, which may not reflect gender identity or sex assigned at birth.

### Supplemental Table 4: Depression cohort descriptive statistics

|  | **Depression code cohort** **(N=149,607)** | **Antidepressant cohort** **(N=215,955)** | **Depression PRS cohort** **(N=61,551)** |
| --- | --- | --- | --- |
| **Sex**^a^ |  |  |  |
| Male | 53223 (35.6%) | 77067 (35.7%) | 27612 (44.9%) |
| Female | 96381 (64.4%) | 138887 (64.3%) | 33939 (55.1%) |
| **Race**^b^ |  |  |  |
| White | 122149 (81.6%) | 177048 (82.0%) | 56729 (92.2%) |
| Black | 16282 (10.9%) | 19931 (9.2%) | 79 (0.1%) |
| Asian | 1365 (0.9%) | 2244 (1.0%) | 183 (0.3%) |
| Multiple | 811 (0.5%) | 1064 (0.5%) | 74 (0.1%) |
| Other | 1419 (0.9%) | 2426 (1.1%) | 313 (0.5%) |
| Unspecified | 7581 (5.1%) | 13242 (6.1%) | 4173 (6.8%) |
| **Age at encounter**^c^ |  |  |  |
| Mean (SD) | 44.2 (19.7) | 45.2 (19.9) | 49.3 (22.6) |
| Median (25%, 75%) | 44.2 (27.7, 59.5) | 45.8 (28.7, 60.9) | 53.8 (33.1, 66.8) |

^a^ Sex refers to EHR-recorded sex, which may not reflect gender identity or sex assigned at birth. Across the depression and antidepressant cohorts, a total of five individuals had an unspecified sex.

^b^ Race refers to EHR-recorded race, which may not reflect self-reported racial identity.

^c^ For the antidepressant and depression cohorts, age at encounter refers to the age at the patient’s earliest antidepressant prescription or depression billing code, respectively. For the depression PRS regression cohort, age at encounter refers to the median age across all of patient’s encounters, EHR-wide.

Abbreviations: PRS, polygenic risk score.

### Supplemental Table 5: Medications included in the antidepressant ClinicScan analysis

| **Generic name** | **Brand names** | **No. patients** |
| --- | --- | --- |
| Sertraline | Zoloft | 38352 |
| Escitalopram | Lexapro | 28990 |
| Citalopram | Celexa | 24303 |
| Amitriptyline | Elavil | 24017 |
| Duloxetine | Cymbalta, Drizalma | 20128 |
| Bupropion | Wellbutrin, Zyban, Forfivo, Budeprion, Aplenzin | 19418 |
| Fluoxetine | Prozac, Sarafem | 18470 |
| Venlafaxine | Effexor | 13203 |
| Mirtazapine | Remeron | 13098 |
| Nortriptyline | Pamelor | 9455 |
| Paroxetine | Paxil, Brisdelle, Pexeva | 8256 |
| Doxepin | Silenor | 2985 |
| Desvenlafaxine | Pristiq | 905 |
| Imipramine | Tofranil | 788 |
| Desipramine | Norpramin | 565 |
| Fluvoxamine | Luvox | 431 |
| Milnacipran | Savella | 420 |
| Vortioxetine | Trintellix, Brintellix | 330 |
| Vilazodone | Viibryd | 256 |
| Clomipramine | Anafranil | 212 |
| Nefazodone | Serzone | 99 |
| Protriptyline | Vivactil | 44 |
| Levomilnacipran | Fetzima | 33 |
| Phenelzine | Nardil | 28 |
| Tranylcypromine | Parnate | 22 |

### Supplemental Table 6: ClinicWAS cohort descriptive statistics

| **Variable** | **Adult ClinicWAS**  **cohort** **(N=55,020)** |
| --- | --- |
| **Sex**^a^ |  |
| Male | 24,176 (43.9%) |
| Female | 30,844 (56.1%) |
| **Race**^b^ |  |
| White | 50,794 (92.3%) |
| Black | 58 (0.1%) |
| Asian | 138 (0.3%) |
| Multiple | 48 (0.1%) |
| Other | 276 (0.5%) |
| Unspecified | 3,706 (6.7%) |
| **Age at Encounter**^c^ |  |
| Mean (SD) | 56.2 (15.8) |
| Median (25%, 75%) | 57.5 (45.4, 68.0) |
| **Elevated genetic risk for coronary heart disease^d^** | 2,645 (4.8%) |

^a^ Sex refers to EHR-recorded sex, which may not reflect gender identity or sex assigned at birth. Across the depression and antidepressant cohorts, a total of five individuals had an unspecified sex.

^b^ Race refers to EHR-recorded race, which may not reflect self-reported racial identity.

^c^ Age is calculated as the median age across all of a patient’s clinical encounters within the health system.

^d^ Elevated genetic risk was defined as having a PRS above the 95^th^ percentile relative to other participants in BioVU.

### Supplemental Table 7: Descriptive statistics for cardiac catheterization cohort

|  | **Cardiac catheterization**  **cohort** **(N=5,120)** |
| --- | --- |
| **Sex**^a^ |  |
| Male | 3,448 (67.3%) |
| Female | 1,672 (32.7%) |
| **Race**^b^ |  |
| White | 5,074 (99.1%) |
| Black | 2 (<0.1%) |
| Asian | 4 (0.1%) |
| Multiple | 6 (0.1%) |
| Other | 15 (0.3%) |
| Unspecified | 19 (0.4%) |
| **Age at Encounter**^c^ |  |
| Mean (SD) | 66.4 (10.7) |
| Median (25%, 75%) | 67.3 (59.3, 74.2) |
| **Elevated genetic risk for coronary heart disease^d^** | 280 (5.1%) |

^a^ Sex refers to EHR-recorded sex, which may not reflect gender identity or sex assigned at birth. Across the depression and antidepressant cohorts, a total of five individuals had an unspecified sex.

^b^ Race refers to EHR-recorded race, which may not reflect self-reported racial identity.

^c^ Age is calculated as the median age across all of a patient’s clinical encounters within the health system.

^d^ Elevated genetic risk was defined as having a PRS above the 95^th^ percentile relative to other participants in BioVU.

### Supplemental Figure 1. Distribution of care sites by the year of the median encounter

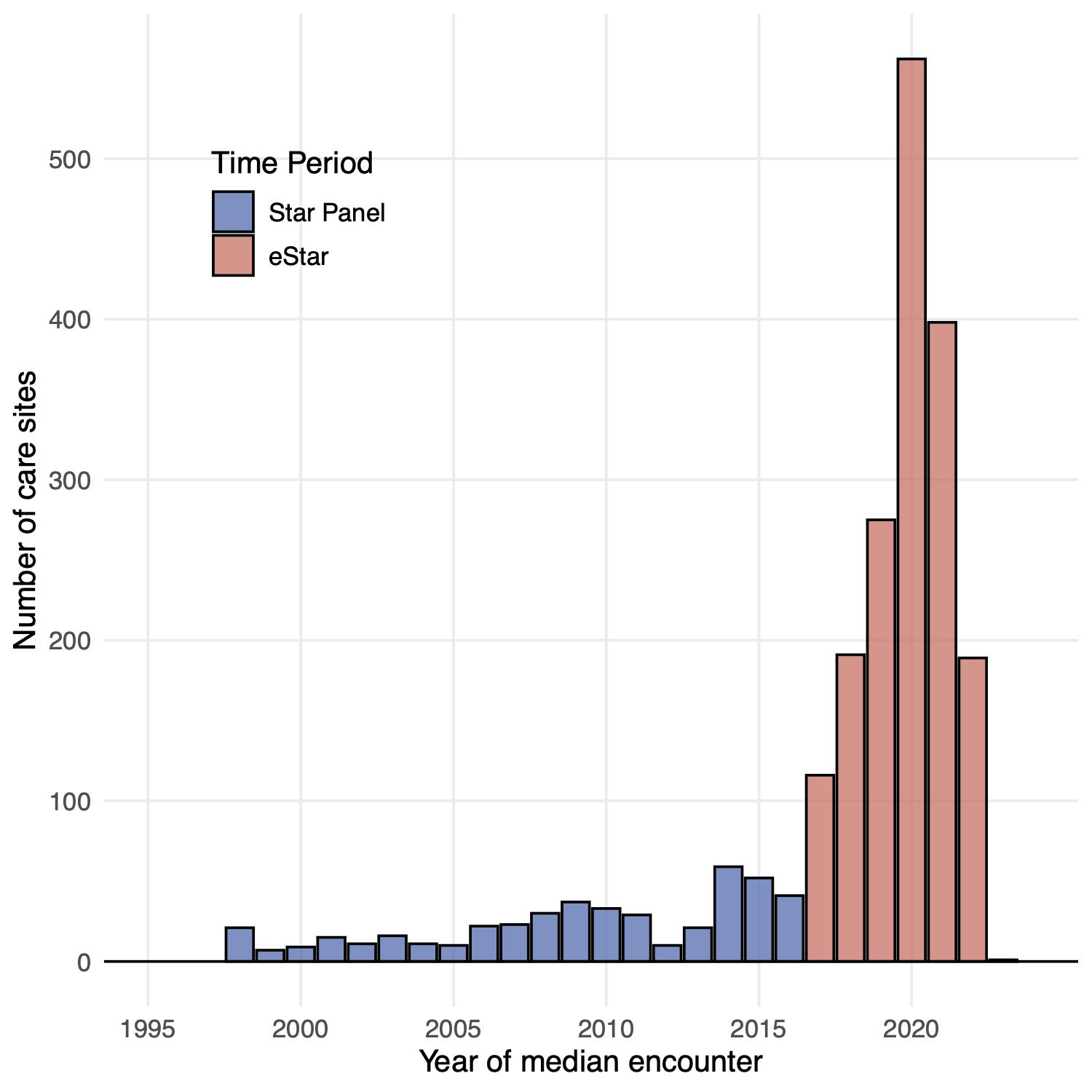

In 2017, VUMC transitioned from a custom designed electronic health record named Star Panel to eStar, a system developed by Epic Systems Corporation (Verona, Wisconsin), leading to an increase in the number of care sites populated in the care_site OMOP CDM table at VUMC.

### Supplemental Figure 2: Co-occurring specialties across care sites.

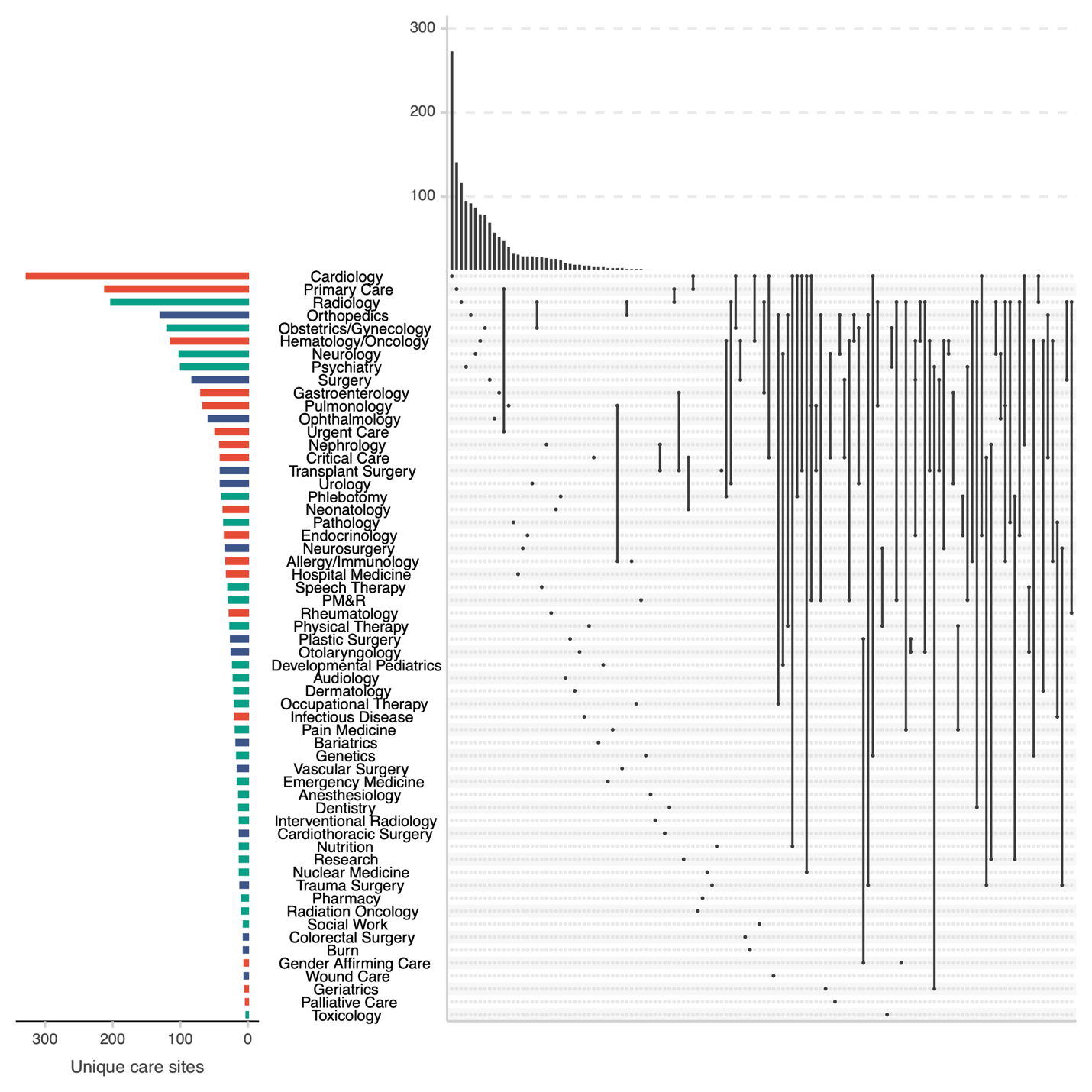

UpSet plot demonstrating patterns of care sites mapping to multiple specialties. Each row represents a clinical specialty, ordered by number of care sites mapping to that specialty, as depicted in the bar plot on the left. Each bar is colored by specialty group: medical (red), surgical (blue), or other (green). Vertical lines between rows represent the intersection of two care sites. The number of care sites in each intersection is shown in the upper histogram.

**Supplemental Figure 3**: Antidepressant prescription ClinicScan plot

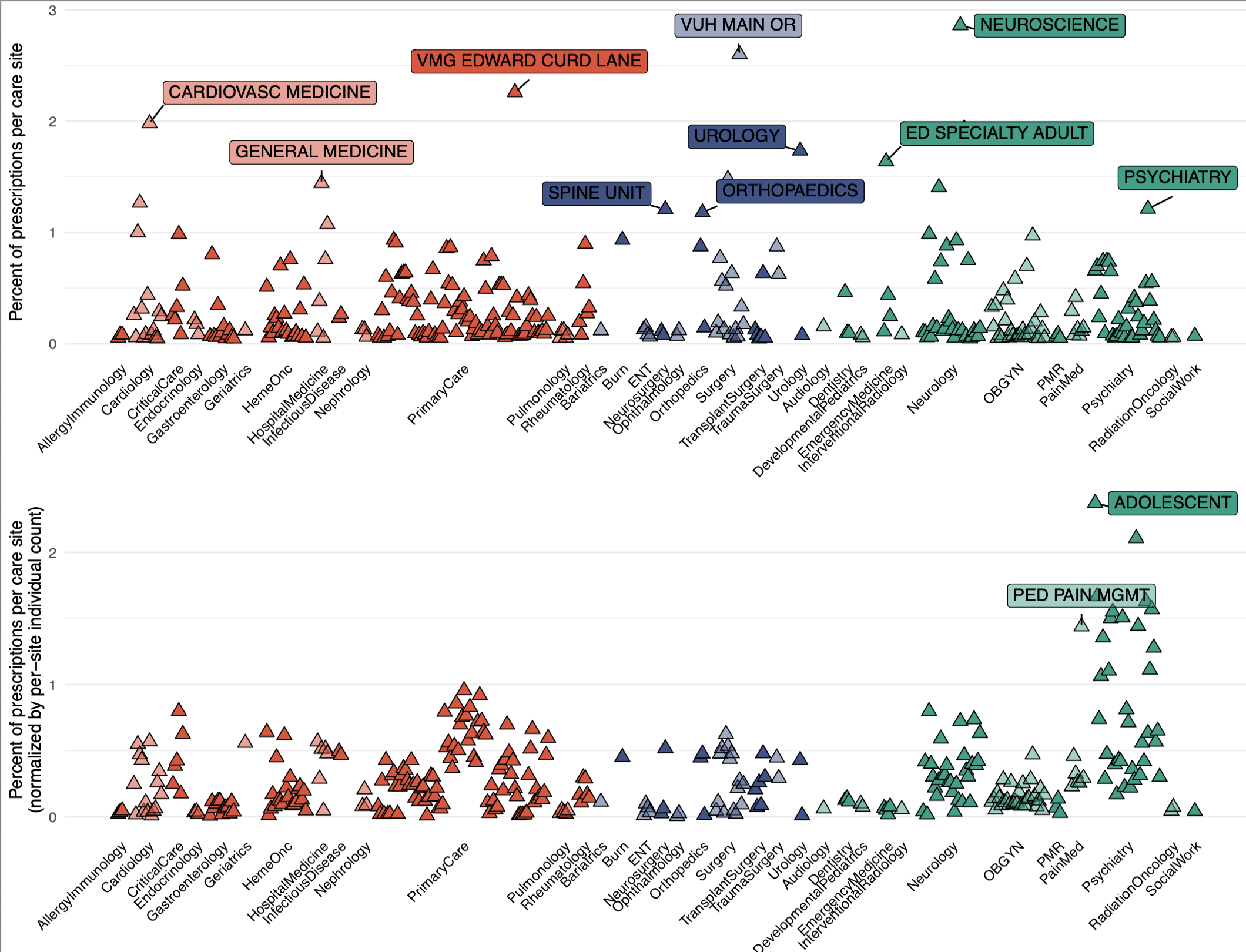

Each point represents the proportion (upper panel) or normalized proportion (lower panel) of antidepressant prescriptions documented in that particular care site. Only the earliest prescription for each patient was considered. Clinical specialties are grouped into medical (red), surgical (blue), and other (green) specialties. Only sites with at least 100 prescriptions are included.

### Supplemental Figure 4. Distribution of the number of CPT codes needed to reach saturation (99% of billing activity) for each care site

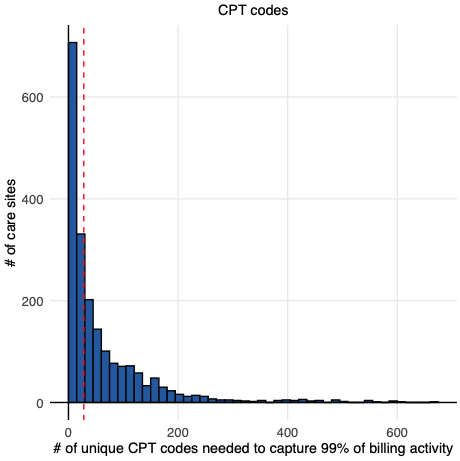

Histogram depicts the distribution of the per-site number of unique CPT codes required to explain 99% of billing activity. For each care site, CPT codes were first ranked according to term frequency (TF) and a running cumulative frequency was calculated for each rank. The saturation point was defined as the rank of the CPT code that met the 99% saturation threshold. The red vertical line depicts the median number of unique CPT codes that explained 99% of billing activity across care sites.

### Supplemental Figure 5. Term frequency (TF) of clinical procedure and service (CPT-4) codes by specialty

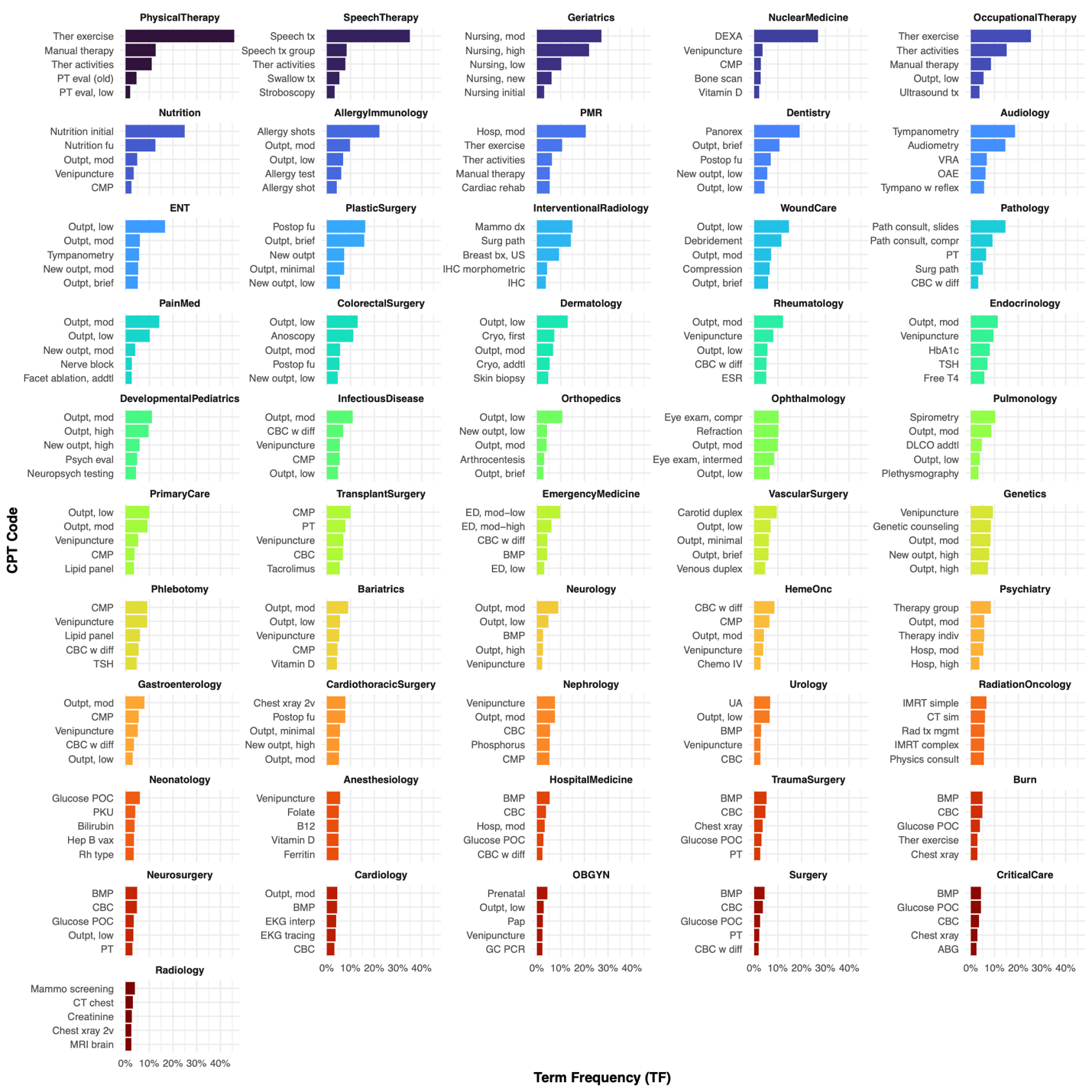
The top 5 procedure and service (CPT-4) billing codes ranked by frequency for each specialty. The frequency of each CPT code out of all CPT code occurrences at that specialty is plotted on the x-axis, while the numeric code is represented on the y-axis. Specialties are sorted according to the TF-IDF score of their highest-ranked code. Non-clinical specialties (pharmacy, social work, research) and specialties with fewer than 1,000 code occurrences (palliative care, gender-affirming care, and toxicology) were excluded. Full rankings of CPT codes for each specialty with descriptions for each CPT code are available in **Supplemental Data 5**.

### Supplemental Figure 6. TF-IDF of clinical procedure and service (CPT-4) codes by specialty

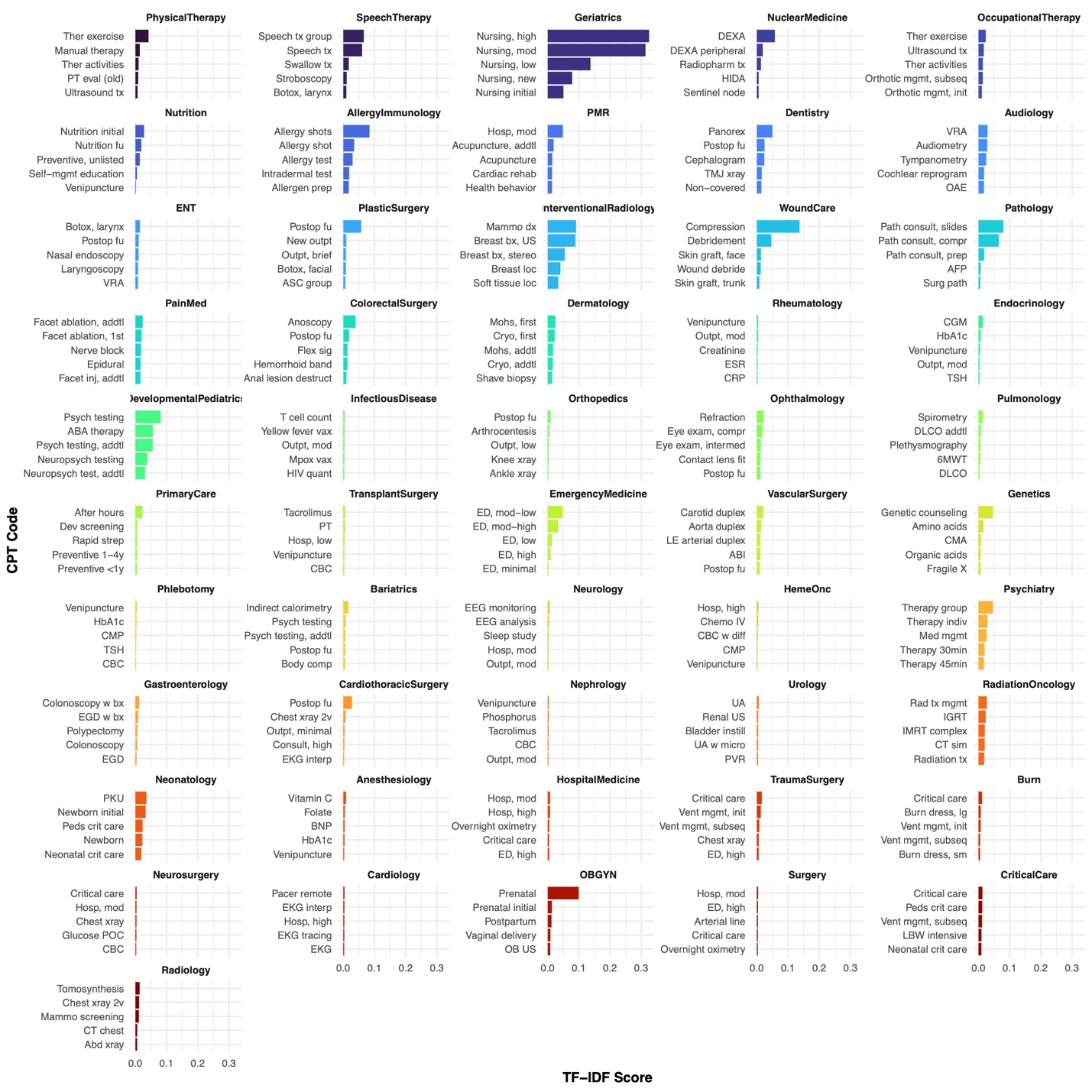
The top 5 procedure and service (CPT-4) billing codes ranked by IDF-weighted TF (TF-IDF) for each specialty. The weighted frequency of each CPT code out of all CPT code occurrences at that specialty is plotted on the x-axis, while the numeric code is represented on the y-axis. Specialties are sorted according to the TF-IDF score of their highest-ranked code. Non-clinical specialties (pharmacy, social work, research) and specialties with fewer than 1,000 code occurrences (palliative care, gender-affirming care, and toxicology) were excluded. Full rankings of CPT codes for each specialty with descriptions for each CPT code are available in **Supplemental Data 5**.
